# Initiation of Warfarin is associated with Decreased Mortality in Patients with Infective Endocarditis: A Population-Based Cohort Study

**DOI:** 10.1101/2023.03.24.23287724

**Authors:** Teddy Tai Loy Lee, Sunny Ching Long Chan, Oscar Hou In Chou, Sharen Lee, Jeffrey Shi Kai Chan, Tong Liu, Carlin Chang, Wing Tak Wong, Gregory Y. H. Lip, Bernard Man Yung Cheung, Abraham Ka-Chung Wai, Gary Tse

## Abstract

**Importance:** The use of warfarin as an anticoagulant to prevent thromboembolism in patients with infective endocarditis (IE) remains controversial due to potentially increased bleeding risks.

**Objective:** This study compared the risks of ischemic stroke, death and bleeding in patients with IE with and without warfarin use.

**Design:** Prospective cohort study.

**Setting:** Population-based.

**Participants:** Patients aged 18 or older and diagnosed with IE in Hong Kong between January 1^st^ 1997 and August 31^st^ 2020 were included. Patients with use of any anticoagulant 30 days before IE diagnosis were excluded. Patients initiated on warfarin within 14 days of IE diagnosis and patients without warfarin use were matched for baseline characteristics using 1:1 propensity score matching.

**Exposure:** Warfarin use within 14 days of IE diagnosis.

**Main outcomes and measures:** Patients were followed up to 90 days for the outcomes of ischemic stroke, all-cause mortality, intracranial hemorrhage, and gastrointestinal bleeding. Cox regression was used to determine hazard ratios (HRs) [95% confidence intervals (CIs)] between treatment groups. Fine-Gray competing risk regression with all-cause mortality as the competing event was performed as a sensitivity analysis. In addition to 90-day analyses, landmark analyses were performed at 30 days of follow-up.

**Results:** The matched cohort consisted of 675 warfarin users (57.0% male, age 59±16 years) and 675 warfarin non-users (53.5% male, age 61±19 years). From Cox regression, warfarin users had a 50% decreased 90-day risk in all-cause mortality (HR: 0.50 [0.39-0.65]), without significantly different 90-day risks of ischemic stroke (HR: 1.04 [0.70-1.53]), intracranial haemorrhage (HR: 1.25 [0.77-2.04]), and gastrointestinal bleeding (HR: 1.04 [0.60-1.78]). Thirty-day landmark analysis showed similar results. Competing risk regression showed significantly higher 30-day cumulative incidence of intracranial haemorrhage in warfarin users (sub-HR: 3.34 [1.34-8.31]), but not at 90-day (sub-HR: 1.63 [0.95-2.81]). Results from Fine-Gray regression were otherwise congruent with those from Cox regression.

**Conclusions and relevance:** In patients with IE, warfarin use initiated within 14 days of IE diagnosis may be associated with significantly decreased risks of mortality but higher risks of intracranial haemorrhage, with similar risks of ischemic stroke and gastrointestinal bleeding, compared with non-use of warfarin with 14 days of IE diagnosis.

**Key points:** *Question:* Is warfarin, initiated within 14 days of a diagnosis of infective endocarditis (IE), efficacious and safe?

*Findings:* In this propensity score-matched, population-based, prospective cohort study from Hong Kong, warfarin use within 14 days of IE diagnosis was associated with a 50% decrease in the risk of all-cause mortality, albeit with higher risk of intracranial haemorrhage, and without significant differences in the risk of ischaemic stroke and gastrointestinal bleeding.

*Meaning:* In patients with IE, warfarin use within 14 days of diagnosis may have mortality benefits, despite increased risks of intracranial haemorrhage.

## Introduction

Infective endocarditis (IE) is an acute infection of the cardiac endothelium, characterized by adherence of platelets and fibrin to the endocardial wall in response to injury, forming a vegetation inducing cardiac damage and development of thromboembolic complications. Despite being a rare condition with an annual incidence of 3-10 cases per 100,000 people ^1, 2^, IE is a challenging condition associated with a high mortality and morbidity, with a reported mortality rate of 30% at 30 days ^3^. Without proper treatment, further platelet aggregation and microbial proliferation allows IE vegetations to grow in size ^4^, subsequently leading to increased risk of septic embolization ^5^. Ischemic stroke, a common and disabling neurological complication caused by an embolized vegetation, has a prevalence of 16.9% ^6^ and is a leading cause of death in IE ^7, 8^.

Though fibrin constitutes the main component in vegetations ^9, 10^, the current 2015 European Society of Cardiology guidelines do not suggest IE per se as an indication for initiating anticoagulation ^11^. However, preclinical studies have shown that anticoagulant use is associated with reduced vegetation size, bacterial load, and inflammation in IE ^12, 13^and may have a role in long-term IE prophylaxis ^14^. Indeed, the role of anticoagulation therapy in IE management is highly controversial due to the associated bleeding risks. Warfarin is a Vitamin K antagonist which inhibits Vitamin K epoxide production of clotting factors and has previously been used to treat valvular vegetations and as prophylaxis against embolic stroke ^15-17^. However, the benefits of prophylaxis must be balanced against bleeding risks, notably intracranial hemorrhage ^18^.

Current data on the efficacy and safety of warfarin therapy in IE are limited, and published clinical studies often have a small numbers of patients. Owing to the lack of evidence, surgical treatment and antibiotics remain the preferred option for reducing the risk of ischemic stroke. Therefore, the present study examined the efficacy in terms of stroke risk reduction, and safety in terms of bleeding risks of warfarin use in a large population-based cohort of patients with IE.

## Methods

This study was been approved by The Chinese University of Hong Kong-New Territories East Cluster Clinical Research Ethics Committee and The University of Hong Kong/Hospital Authority Hong Kong West Cluster Institutional Review Board. The need for informed consent was waived owing to the retrospective use of deidentified patient data in this study. Patient data was obtained through the Clinical Data Analysis and Reporting System (CDARS), a territory-wide electronic healthcare database managed by the Hong Kong Hospital Authority, which serves an estimated 90% of the population in Hong Kong. CDARS has previously been used extensively to conduct large population-based studies ^19, 20^, including those on anticoagulation use ^21, 22^.

### Study Cohort

All patients aged 18 or above with a diagnosis of IE who attended any public hospitals between January 1^st^, 1997 to August 31^st^, 2020 were identified from CDARS. The date of first diagnosis of IE was defined as the index date. Patients who received warfarin prescriptions within 14 days since the index date were considered warfarin users. To select new users only, patients who received warfarin or other anticoagulant medications (rivaroxaban, dabigatran, apixaban, edoxaban, enoxaparin, fondaparinux or heparin) within an entry period 30 days prior to the index date were excluded.

### Outcomes

To study the efficacy and safety of warfarin use, study outcomes included 90-day risks of embolic stroke and all-cause mortality (efficacy), and intracranial hemorrhage and gastrointestinal bleeding (safety). An *a priori* landmark analysis was done at 30 days to compare the 30-day risks. Details of International Classification of Diseases, Ninth Revision (ICD-9) codes used to define outcomes are described in **Supplementary Table 1**. Outcomes were followed up until the occurrence of outcome, death, or until 90 days after IE diagnosis, whichever earlier.

### Covariates

We traced patient records on CDARS prior to the index date and collected patient information including age at index date, sex, comorbidities (hypertension, atrial fibrillation, heart failure, diabetes mellitus, ischemic heart disease, chronic kidney disease, vascular disease), history of valvular replacement, and recent medication use, including drugs related to bleeding risk (ACE inhibitors, angiotensin receptor blockers, beta blockers, nonsteroidal anti-inflammatory drugs [NSAIDs], histamine type 2 receptor antagonists [H2RAs], proton pump inhibitors [PPIs], selective serotonin reuptake inhibitors [SSRIs]), and pathogens identified from blood culture (staphylococci, streptococci, enterococci, HACEK group) taken during IE admission. We also traced the number of patients who received direct oral anticoagulants (DOACs), heparin (fondaparinux, enoxaparin, heparin), and cardiac surgery (ICD-9-CM Procedure codes 35-39) within 14 days since the index date. ICD-9 codes used to identify comorbidities are described in **Supplementary Table 1**.

### Statistical Analysis

This study tested the hypothesis that when compared to non-usage, warfarin usage is associated with different risks of efficacy and safety study outcomes of ischemic stroke, all-cause mortality, intracranial hemorrhage and gastrointestinal bleeding. To account for differences between groups in baseline characteristics due to a lack of randomization, we used propensity score matching on a 1:1 ratio using the nearest-neighbour matching algorithm to match warfarin users with non-users by the aforementioned covariates. A caliper width of 0.2 was chosen as it is considered optimal for propensity score matching ^23^. Standardized mean differences (SMD), the differences in means over the pooled standard deviation (SD) assessed balance of categorical covariates; the variance ratio (VR), which is the ratio of variance between the treatment and control groups assessed balance of both categorical and continuous covariates. Characteristics with an SMD of <0.1 or VR between the range of 0.5 and 2.0 were considered balanced. Descriptive statistics were expressed as mean ± SD and count (percentage [%]) as appropriate.

Result estimates were expressed in terms of hazard ratios (HRs) with 95% confidence intervals (CIs) using a univariate Cox proportional hazards model. The proportional hazards assumption of the model was tested by performing the Schoenfeld proportionality test; the results indicated that the assumption was met. Kaplan-Meier curves were plotted against the time-to-event, beginning from the date of IE diagnosis stratified by either warfarin use or no warfarin use for the main results, and beginning from the date of warfarin initiation and stratified by either early or late initiation of warfarin for analysis of warfarin timing. The log-rank test was performed to investigate the statistical significance of differences in survival between comparator groups.

An *a priori* subgroup analysis was performed: 90-day risks of the efficacy outcomes in patients who were initiated on warfarin early (≤7 days) were compared to those with late initiation (within 8 to 14 days) to explore the effect of initiation timing within warfarin users; in this analysis, the start date of follow-up was defined as the date of warfarin initiation instead of date of IE diagnosis. As there were substantial imbalances between treatment arms in the respective proportions of patients who received cardiac surgery and heparin within 14 days of the index day, two *post hoc* subgroup analyses were added, where patients who received cardiac surgery and heparin were excluded separately. Furthermore, two *a priori* sensitivity analyses were conducted. First, the period in which warfarin was initiated was restricted from within 14 days to within 7 days of the index date. Second, a competing risk regression was performed for ischaemic stroke, intracranial haemorrhage, and gastrointestinal bleeding using the Fine and Gray sub-distribution model, with all-cause mortality as the competing event, and with sub-hazard ratios (SHRs) and the corresponding 95% CIs as summary statistics. Aalen-Johansen cause-specific cumulative incidence curves were additionally plotted for these outcomes to account for competing risks.

All analyses were conducted using R (version 1.4.1717) or Stata (version 16.1). All p-values were two-tailed and considered statistically significant when P < 0.05.

## Results

### Patient characteristics

The patient flow is shown in **Figure 1**. In total, 7,054 patients with a diagnosis of IE were identified from CDARS. After excluding patients with previous anticoagulant use, under 18 years old, and those with use of warfarin after 14 days since IE diagnosis, the study cohort consisted of 5,477 patients, where 734 patients received warfarin (56.7% male, mean baseline age: 60±16 years old) and 4743 without warfarin use (66.5% male, mean baseline age: 58±20 years old). All patients completed 90-day follow up or had 90-day mortality.

**Figure 1:**
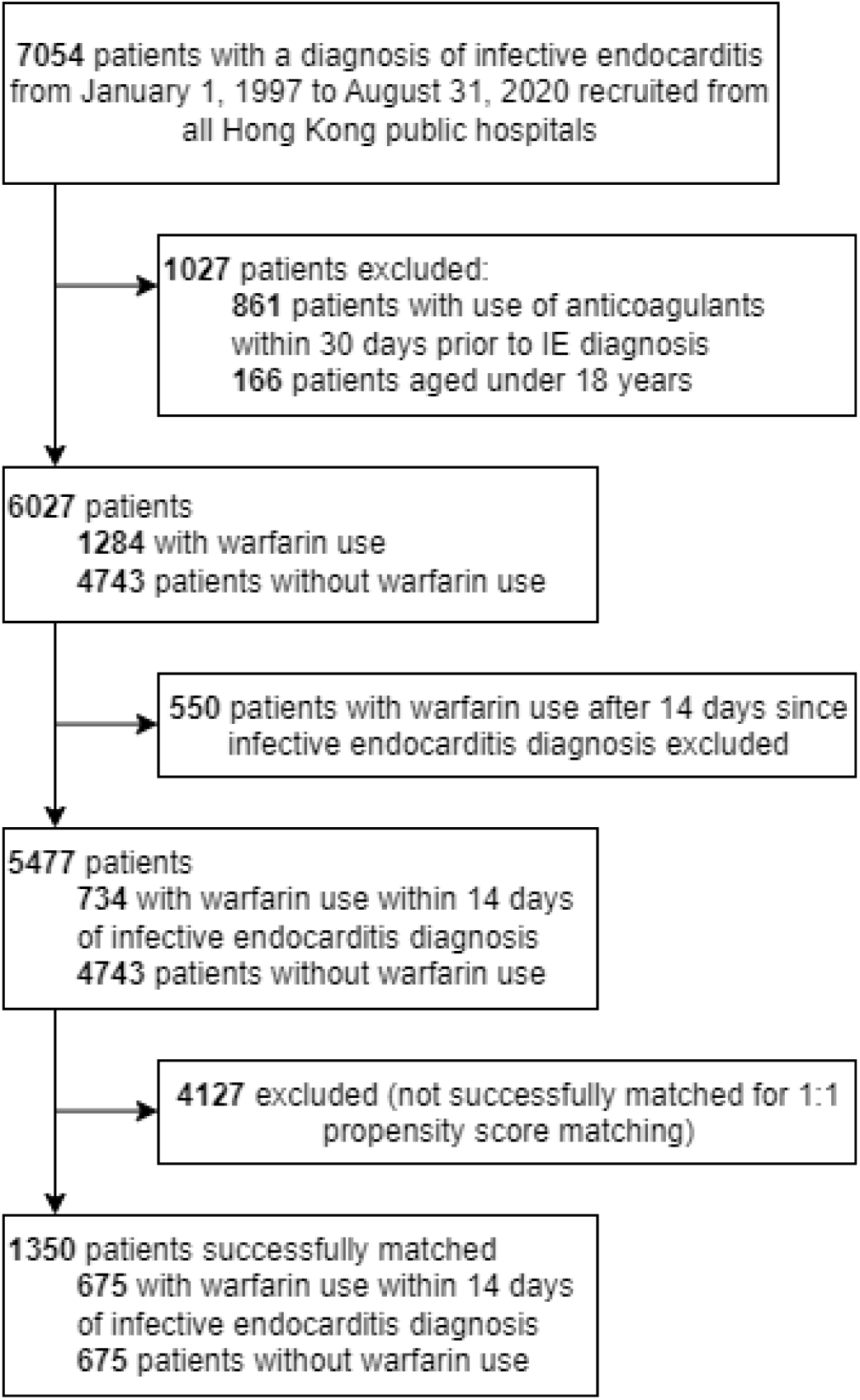
Study flow chart.

After 1:1 propensity score matching, the final study cohort consisted of 675 warfarin users and 675 without warfarin use. The baseline characteristics of the study population are shown in **Table 1**. A Love plot summarizing covariate balance before and after propensity score matching is shown in **Supplementary Figure 1**. All SMD values were <0.1 and VR values within 0.5-2.0, indicating good balance between warfarin users and those without warfarin use; the only exception being age, which had a SMD of 0.13. After matching, 13 (1.9%) warfarin users and 23 (3.4%) patients without warfarin use received DOACs, while 435 (64.4%) warfarin users 68 (10.1%) patients without warfarin use received heparin. 162 (24.0%) warfarin users compared to 59 (8.7%) patients without warfarin use received cardiac surgery within 14 days of IE diagnosis.

**Table 1:** Characteristics of patients with infective endocarditis before and after 1:1 propensity score matching.

| Characteristics | Before matching |  | After matching |  |  |  |
| --- | --- | --- | --- | --- | --- | --- |
|  | Warfarin use (N=734) | No warfarin use (N=4743) | Warfarin use (N=675) | No warfarin use (N=675) | SMD | VR |
| **Demographics** |  |  |  |  |  |  |
| Male, N (%) | 416 (56.7) | 3156 (66.5) | 385 (57.0) | 361 (53.5) | 0.04 | - |
| Baseline age, years | 59.63±15.89 | 58.12±19.64 | 59.36±16.25 | 61.45±18.74 | 0.13 | 0.75 |
| **Comorbidities** |  |  |  |  |  |  |
| Charlson Comorbidity Index | 2.44±2.27 | 2.08±2.39 | 2.37±2.29 | 2.51±2.38 | 0.06 | 0.93 |
| Hypertension, N (%) | 137 (18.7) | 828 (17.5) | 126 (18.7) | 137 (20.3) | 0.02 | - |
| Atrial fibrillation, N (%) | 253 (34.5) | 329 (6.9) | 206 (30.5) | 213 (31.6) | 0.01 | - |
| Heart failure, N(%) | 229 (31.2) | 522 (11.0) | 187 (27.7) | 202 (29.9) | 0.02 | - |
| Diabetes mellitus, N (%) | 83 (11.3) | 545 (11.5) | 78 (11.6) | 85 (12.6) | 0.01 | - |
| Intracranial hemorrhage, N (%) | 25 (3.4) | 108 (2.3) | 21 (3.1) | 24 (3.6) | <0.01 | - |
| Chronic kidney disease, N (%) | 28 (3.8) | 295 (6.2) | 28 (4.1) | 25 (3.7) | <0.01 | - |
| Vascular disease, N (%) | 8 (1.1) | 36 (0.8) | 6 (0.9) | 7 (1.0) | <0.01 | - |
| Valvular replacement, N (%) | 107 (14.6) | 43 (0.9) | 52 (7.7) | 39 (5.8) | 0.02 | - |
| **Medications** |  |  |  |  |  |  |
| ACE inhibitors, N (%) | 151 (20.6) | 473 (10.0) | 130 (19.3) | 147 (21.8) | 0.02 | - |
| Angiotensin receptor blockers, N (%) | 38 (5.2) | 124 (2.6) | 33 (4.9) | 32 (4.7) | <0.01 | - |
| Beta blockers, N (%) | 149 (20.3) | 510 (10.8) | 132 (19.6) | 135 (20.0) | <0.01 | - |
| NSAIDs, N (%) | 168 (22.9) | 847 (17.9) | 157 (23.3) | 170 (25.2) | 0.02 | - |
| H2 receptor antagonists, N (%) | 147 (20.0) | 664 (14.0) | 131 (19.4) | 138 (20.4) | 0.01 | - |
| Proton pump inhibitors, N (%) | 136 (18.5) | 600 (12.7) | 116 (17.2) | 118 (17.5) | <0.01 | - |
| Selective serotonin reuptake inhibitors, N (%) | 8 (1.1) | 47 (1.0) | 8 (1.2) | 6 (0.9) | <0.01 | - |
| **Blood culture results** |  |  |  |  |  |  |
| Staphylococci, N (%) | 87 (11.9) | 1023 (21.6) | 73 (10.8) | 69 (10.2) | <0.01 | - |
| Streptococci, N (%) | 68 (9.3) | 681 (14.4) | 58 (8.6) | 52 (7.7) | <0.01 | - |
| Enterococci, N (%) | 12 (1.6) | 112 (2.4) | 8 (1.2) | 11 (1.6) | <0.01 | - |
| HACEK group, N (%) | 1 (0.1) | 33 (0.7) | 1 (0.1) | 3 (0.4) | <0.01 | - |
| **Outcomes** |  |  |  |  |  |  |
| Ischemic stroke, N (%) | 57 (7.8) | 314 (6.6) | 52 (7.7) | 50 (7.4) | - | - |
| All-cause mortality, N (%) | 93 (12.7) | 1246 (26.3) | 83 (12.3) | 193 (28.6) | - | - |
| Intracranial hemorrhage, N (%) | 39 (5.3) | 184 (3.9) | 36 (5.3) | 29 (4.3) | - | - |
| Gastrointestinal bleeding, N (%) | 30 (4.1) | 146 (3.1) | 27 (4.0) | 26 (3.9) | - | - |
| ***Treatments received within 14 days after IE diagnosis*** |  |  |  |  |  |  |
| Direct oral anticoagulants, N (%) | 13 (1.8) | 47 (1.0) | 13 (1.9) | 23 (3.4) | - | - |
| Heparin, N (%) | 466 (63.5) | 368 (7.8) | 435 (64.4) | 68 (10.1) | - | - |
| Cardiac surgery, N (%) | 171 (23.3) | 453 (9.6) | 162 (24.0) | 59 (8.7) | - | - |

Continuous variables were expressed as mean ± standard deviation. SMD and VR were only displayed for variables considered in propensity score matching. SMD <0.1 / VR >0.5 & 2.0 indicated good balance in matching.
ACE, Angiotensin-converting enzyme; IE: Infective endocarditis; NSAID: Nonsteroidal anti-inflammatory drugs; SMD: Standardized mean difference; VR: Variance ratio.

The ten most common causes of death were described in **Supplementary Table 2**. The most common causes of death in warfarin users were IE (7.0%), followed by pneumonia (2.2%), sepsis (1.2%), heart failure (0.9%), and rheumatic heart disease (0.6%); while in patients without warfarin use, the most common causes of death were IE (2.5%), followed by pneumonia (1.2%), stroke (0.9%), rheumatic heart disease (0.6%), and intracranial hemorrhage (0.6%). 2 (0.3%) patients without warfarin use and 6 (0.9%) patients with warfarin use died of stroke, while 6 (0.8%) patients without warfarin use and 7 (1.0%) patients with warfarin use died of intracranial, intracerebral or subarachnoid hemorrhage **(Supplementary Table 3)**.

### Main 90- and 30-day analytic results

The main analytic results are presented in **Table 2**, and Kaplan-Meier plots are shown in **Figure 2**. While warfarin use was not associated with a significantly different 90-day risk of ischemic stroke (HR: 1.04 [95% CI, 0.70-1.53], log-rank p=0.86), it was associated with a 50% decrease in the 90-day risk of all-cause mortality (HR: 0.50 [0.39-0.65], log-rank p<0.0001). For the safety outcomes, warfarin use was not associated with significantly different 90-day risks of gastrointestinal bleeding (HR 1.04 [0.60-1.78], log-rank p=0.90), although a nonsignificant trend for intracranial hemorrhage (HR: 1.25 [0.77-2.04]; log-rank p=0.37) may be present.

**Table 2:**
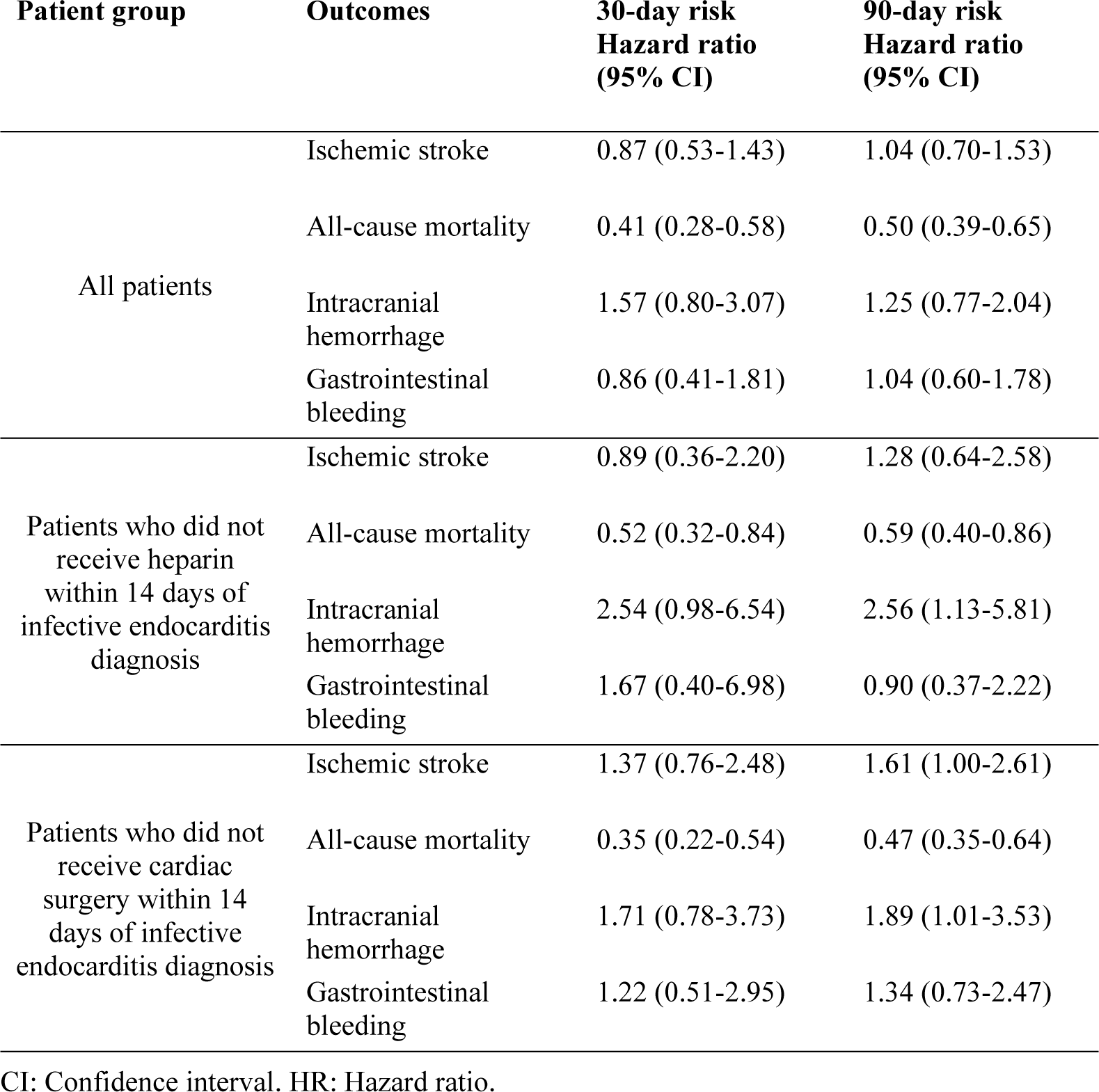
Main and subgroup analyses of study outcomes.

**Figure 2:**
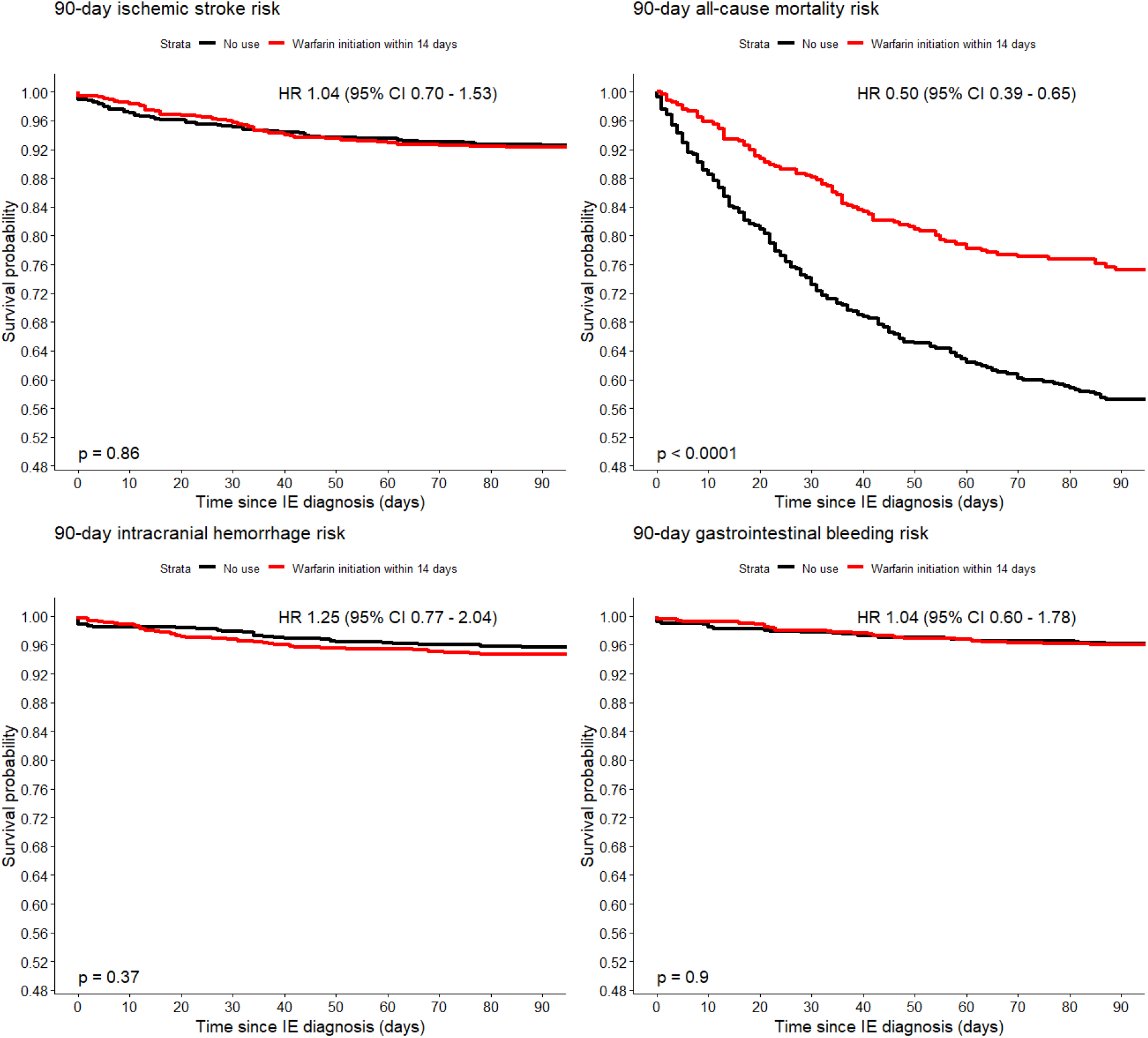
Kaplan-Meier curves of cumulative freedom from the study outcomes, stratified by warfarin use. CI, confidence interval. HR, hazard ratio

Results of the 30-day landmark analysis for ischemic stroke (HR: 0.87 [0.53-1.43]), all-cause mortality (HR: 0.41 [0.28-0.58]), intracranial hemorrhage (HR: 1.57 [0.80-3.07]) and gastrointestinal bleeding (HR: 0.86 [0.41-1.81]) were consistent with those of the main, 90-day analysis.

### Subgroup analysis

Results of subgroup analyses are shown in **Table 2** as well. In the *a priori* subgroup analysis exploring the effects of the timing of warfarin initiation, warfarin initiation between 8-14 days of IE diagnosis (delayed use; N=152) was not associated with significantly different 90-day risks of ischemic stroke (HR: 1.53 [0.82-2.86]) or all-cause mortality (HR: 1.29 [0.78-2.14]) compared with early use (warfarin initiation within the seven days of IE diagnosis; N=523; **Figure 3**). Landmark analysis at 30 days showed consistent results (**Supplementary Table 4**).

**Figure 3:**
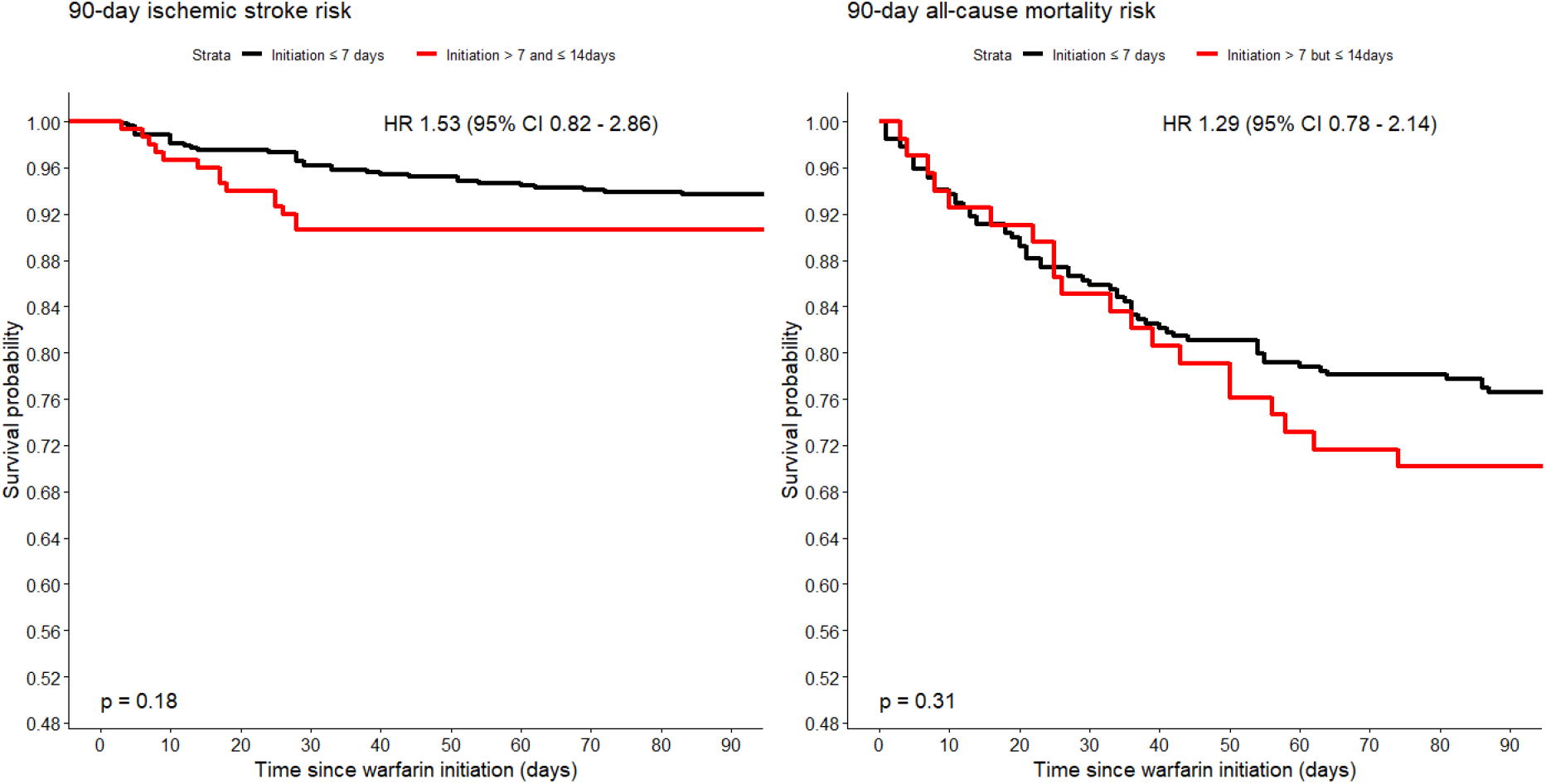
Kaplan-Meier survival curves of study outcomes, stratified by the timing of warfarin initiation. CI, confidence interval. HR, hazard ratio.

In the first *post hoc* subgroup analysis excluding patients who received heparin within 14 days of IE diagnosis, warfarin use was associated with significantly lower 90-day risk of all-cause mortality (HR: 0.59 [0.40-0.86]), but also significantly higher 90-day risk of intracranial haemorrhage (HR: 2.56 [1.13-5.81]; **Supplementary Figure 2**); the 90-day risk of intracranial haemorrhage and gastrointestinal bleeding were not significantly different between treatment arms. Landmark analysis at 30 days showed largely consistent results, except for the nonsignificant trend in the 30-day risk of intracranial haemorrhage between treatment arms (HR: 2.54 [0.98-6.54]).

In the second *post hoc* subgroup analysis excluding patients who received cardiac surgery within 14 days of IE diagnosis, warfarin use was again associated with significantly lower 90-day risk of all-cause mortality (HR: 0.47 [0.35-0.64]), but also significantly higher 90-day risk of intracranial haemorrhage (HR: 1.89 [1.01-3.53]; **Supplementary Figure 3**); the 90-day risk of intracranial haemorrhage and gastrointestinal bleeding were not significantly different between treatment arms. Landmark analysis at 30 days showed largely consistent results, except the 30-day risk of intracranial haemorrhage was not significantly different between treatment arms (HR: 1.71 [0.78-3.73]).

### Sensitivity analysis

Results of sensitivity analyses were summarized in **Table 3**. When only patients with warfarin use within seven days of IE diagnosis were included in the warfarin arm (N=523), warfarin use was associated with significantly lower 90-day risk of all-cause mortality (HR: 0.51 [0.38-0.67]), but not significantly different 90-day risks of ischemic stroke (HR: 0.81 [0.54-1.23]), intracranial hemorrhage (HR: 0.75 [0.45-1.26]), nor gastrointestinal bleeding (HR: 0.56 [0.30-1.03]).

**Table 3:**
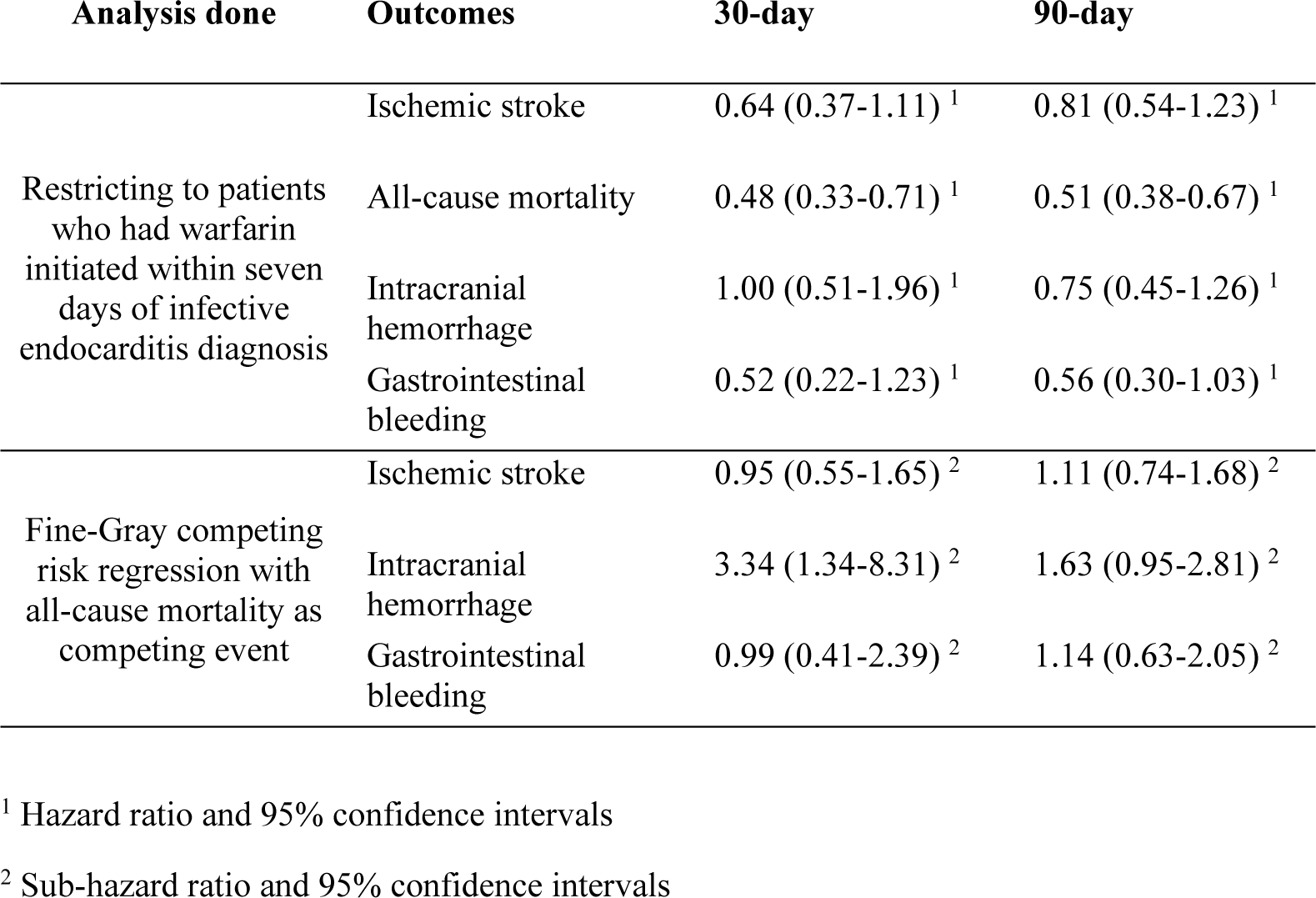
Results of sensitivity analyses.

In the Fine and Gray competing risk regression with all-cause mortality as competing events, warfarin users had significantly higher 30-day cumulative incidence of intracranial haemorrhage (SHR 3.34 [1.34-8.31]; **Supplementary Figure 4**), but the 90-day cumulative incidence was not significantly different between groups (SHR 1.63 [0.95-2.81]). The cumulative incidences of iscahemic stroke (**Supplementary Figure 5**) and gastrointestinal bleeding (**Supplementary Figure 6**) were not significantly different between groups at both 30 and 90 days.

## Discussion

To the best of the authors’ knowledge, this is the first population-based study on warfarin use in IE patients. In this study, warfarin use in patients with IE was associated with a significantly lower risk of mortality, without significantly different risks of ischemic stroke. However, warfarin use may be associated with significantly higher risk of intracranial haemorrhage, despite a lack of significant differences in the risk of other bleeding events.

The most important finding of this study was that in patients with IE, warfarin use was associated with lower mortality risks at both 30 and 90 days. The reduction of clotting factors and reduced platelet aggregation through warfarin use may slow the local proliferation of vegetation, reducing valvular destruction and the extent of the infection, leading to improved prognosis. Clinically, these properties of warfarin are most leveraged for ischaemic stroke prevention. In this study, however, the mortality benefit observed for warfarin use was clearly not driven by ischaemic stroke risk reduction, as warfarin use was not associated with significantly different risk of ischaemic stroke. This contrasts with two studies focused on left-sided staphylococcus aureus IE, where anticoagulation was found to reduce stroke risk ^24, 25^. For example, Rasmussen and colleagues described in a study of 175 IE patients that anticoagulant use, defined as use of either coumadin or high-dose low molecular weight heparin, was associated with a fourfold decrease in stroke risk (adjusted odds ratio [aOR]: 0.27), but most patients who received anticoagulants had prosthetic valves (73%) compared to those without anticoagulant use (8%) ^24^. In another cohort study of 587 IE episodes, warfarin use was associated with a lower risk of cerebrovascular complications (aOR: 0.26 [95% CI, 0.07-0.94]), where 38% of patients with warfarin use and 12% patients without warfarin use had atrial fibrillation on admission ^25^. Nonetheless, both studies included patients who already had continuous use of anticoagulants prior to IE admission. As patients already on treatment may have stably reached the therapeutic international normalized ratio (INR) range, the baseline risk for stroke of the anticoagulant group may be lower at the time of IE diagnosis. Instead, to ensure a new-user design, we excluded patients with prior use of any anticoagulant within 30 days before IE diagnosis, and balanced the coagulability status of treatment groups by matching with medications that influence coagulability, which may explain the lack of significant benefit among new users in reducing ischemic stroke risk compared to previous studies.

The mortality benefit may be driven by a reduction in the size of vegetation, as suggested by previous studies ^15, 24^. As echocardiography data were not available, this could not be verified in our study. It is also unclear whether effects on the size of vegetation was affected by the causative organism. Another potential driver of the observed associations may be differences in the risks of thromboembolic complications such as septic pulmonary embolism ^26^. Septic pulmonary emboli pose high risk to patients as it can cause subsequent respiratory failure requiring mechanical ventilation and prolonging hospital stay ^27^. Though anticoagulation has a well-established role for prophylaxis of noninfective pulmonary embolism ^28^, it is not used for active treatment of septic pulmonary emboli due to the increased bleeding risk in area of the infected emboli ^5^. Nevertheless, treatment of septic pulmonary emboli using anticoagulation has been described in small cohort studies ^29, 30^. Further research on the potential effects of warfarin on vegetation size and thromboembolic complications and the prognostic implications of any such effect is warranted.

The use of antiplatelet therapy in reducing mortality or embolic risk in IE is similarly controversial. In a retrospective cohort study of 600 IE patients, patients with prior use of aspirin, dipyridamole, clopidogrel or ticlopidine had a lower risk of stroke compared to control, but not significantly different mortality ^31^. In a randomized controlled trial of 115 IE patients, treatment with aspirin 325mg/day did not significantly reduce the risk of embolic stroke nor in-hospital mortality, instead increasing the risk of bleeding ^32^; also, this trial excluded patients with previous antiplatelet use, similar to our study. Interestingly, low-dose aspirin, compared to high-dose aspirin, was more effective at reducing bacterial density and vegetation weight ^33^. Furthermore, a recent cohort study involving 34 IE patients compared long term use of antiplatelets or anticoagulants and patients without the use of either medication class. Embolic events occurred in 30% of patients receiving treatment and 7.1% not receiving treatment, with similar mortality risk between both groups ^34^. The study found a lower number of bleeding events in the group without antiplatelet/anticoagulant use, in agreement with the subgroup analysis in the current study where patients who received heparin within 14 days of IE diagnosis were excluded. In local clinical practice, the vast majority of admitted IE patients are started on warfarin instead of DOACs due to valvular nature of the disease; therefore, it was not possible to conduct separate analyses investigating DOAC use on the study outcomes. It remains to be elucidated whether or not DOACs are similarly associated with a decreased risk of mortality in the context of IE. Overall, there is a lack of consensus on the role of antithrombotics in general in the management of IE, be it anticoagulants or antiplatelets, and the use of systemic antibiotics remains the preferred strategy to reduce the risks of mortality and septic embolization ^11, 35, 36^.

Previous evidence on anticoagulation in IE is mostly based on case reports or non-representative observational studies, which were limited by small sample sizes and sampling bias. Nonetheless, a placebo-controlled trial on this study topic is difficult to organize given the potential risk of bleeding in warfarin use. The present study attempted to minimize indication bias for warfarin prescription by propensity score matching with potential confounders such as prosthetic valve replacement, CHA_2_DS_2_-VASc score and atrial fibrillation. The population-based nature of the study also meant that the results are widely generalizable, at least to other developed Asian cities. Moreover, we performed a number of subgroup and sensitivity analyses to ensure robustness of our analyses, observing mostly consistent findings in patients who did not receive cardiac surgery or heparin, and in both patients with early and late warfarin use. The subgroup analysis of patients who did not receive cardiac surgery was especially important, as the implantation of mechanical prosthetic valves could have been the primary indication of warfarin in some patients. By demonstrating consistent results in patients who did not receive cardiac surgery, the associations between warfarin use and lower mortality risk was likely to be explained by factors other than warfarin being a surrogate for surgical treatment. Nonetheless, it must be stressed that clinicians should be aware that warfarin use was likely associated with higher risk of intracranial haemorrhage, as shown in the two *post hoc* subgroup analyses of patients and in the competing risk regression. Although mortality benefit was shown despite such detrimental associations with bleeding risks, more work is needed to carefully delineate the risk-benefit balance in using warfarin in patients with IE, especially with the consideration of morbidity in addition to mortality, before more definitive recommendations can be made for clinical practice.

### Limitations

The present study has several limitations. First, there was incomplete baseline INR data of patients within the treatment group and the coagulability status of study patients before initiation on warfarin is unknown. This was addressed by excluding patients with use of anticoagulants within 30 days before IE diagnosis in both treatment groups. Second, this study did not account for the treatment duration, interruption or discontinuation of therapy in warfarin users, which may influence ischemic stroke or mortality risk. Major bleeding events such as intracranial hemorrhage and gastrointestinal bleeding may prompt interruption of anticoagulant therapy ^11^, which, if abrupt, may lead to a sharp drop in INR ^37^, in turn increasing the risk of ischemic stroke. Third, this was an observational study; owing to limitations of the CDARS database, echocardiographic findings such as the size of vegetation, valvular destruction, and perivalvar abscesses were not coded in the system and therefore could not be explored. Fourth, although a number of baseline characteristics were used for propensity score matching, the observational nature of this study meant that unobserved and residual confounders cannot be completely eliminated. Nonetheless, we believe the baseline characteristics considered should be pragmatically sufficient as a representation of the overall comorbid status of the included patients. Lastly, as all diagnoses and outcomes were ascertained using ICD codes, miscoding may be possible. Nonetheless, CDARS and the linked Hong Kong Death Registry have been used extensively in peer-reviewed studies ^38-40^, and all coding were performed by clinicians independent of the authors.

## Conclusions

In patients with IE, warfarin use may be associated with decreased risks of mortality compared with non-use of warfarin, with a non-significant difference in the risk of ischaemic stroke and a higher risk of intracranial haemorrhage. Larger studies are warranted to confirm these findings and delineate the underlying mechanisms.

## Data Availability

The data used for analysis in this manuscript are available from the corresponding author upon reasonable request.

## Non-standard Abbreviations and Acronyms

ACE: Angiotensin-converting enzyme
CDARS: Clinical data analysis and reporting system
CI: Confidence interval
DOAC: Direct oral anticoagulant
HACEK: Hemophilus species, Aggregatibacter actinomycetemcomitans, Cardiobacterium hominis, Eikenella corrodens, Kingella kingae
HR: Hazard ratio
H2RA: Histamine type 2 receptor antagonist
ICD-9: International Classification of Diseases, Ninth Revision
INR: International normalized ratio
IE: Infective endocarditis
NSAID: Nonsteroidal anti-inflammatory drug
OR: Odds ratio
PPI: Proton pump inhibitor
SD: Standard deviation
SHR: Sub-hazard ratio
SMD: Standardized mean difference
VR: Variance ratio

## Acknowledgements

None.

## Sources of Funding

None.

## Disclosures

None.

## Supplemental Material

Tables S1-S3

Figures S1-S6

## Notes

### Competing Interest Statement

The authors have declared no competing interest.

### Clinical Trial

this is not a clinical trial

### Author Declarations

This study was been approved by The Chinese University of Hong Kong-New Territories East Cluster Clinical Research Ethics Committee and The University of Hong Kong/Hospital Authority Hong Kong West Cluster Institutional Review Board.

